# Children with COVID-19 like symptoms in Italian Pediatric Surgeries: the dark side of the coin

**DOI:** 10.1101/2020.07.27.20149757

**Authors:** Gianfranco Trapani, Vassilios Fanos, Enrico Bertino, Giulia Maiocco, Osama Al Jamal, Michele Fiore, Vincenzo Bembo, Domenico Careddu, Lando Barberio, Luisella Zanino, Giuseppe Verlato

**Author notes:** Corresponding author Dr. Giulia Maiocco, (GM).

## Abstract

**Background:** Symptoms of SARS-CoV-2 infection in children are nonspecific and shared with other common acute viral illnesses (fever, respiratory or gastrointestinal symptoms, and cutaneous signs), thus making clinical differential diagnosis tricky. In Italy, first line management of pediatric care is handed over to Primary Care Pediatricians (PCPs), who were not allowed to directly perform diagnostic tests during the recent COVID-19 outbreak. Without a confirmatory diagnosis, PCPs could only collect information on “COVID-19 like symptoms” rather than identify typical COVID-19 symptoms.

**Aim:** To evaluate the prevalence of COVID-19 like symptoms in outpatient children, during Italian lockdown. To provide PCPs a risk score to be used in clinical practice during the differential diagnosis process.

**Methods:** A survey was submitted to 50 PCPs (assisting 47,500 children) from 7 different Italian regions between the 4^th^ of March and the 23^rd^ of May 2020 (total and partial lockdown period). COVID-19 like symptoms in the assisted children were recorded, as well as presence of confirmed/suspected cases in children’s families, which was taken as proxy of COVID-19. Multivariable logistic regression was accomplished to estimate the risk of having suspected/confirmed cases in families, considering symptoms as potential determinants.

**Results:** 2,300 children (4.8% of overall survey population) fell ill with COVID-19 like symptoms, 3.1% and 1.7% during total and partial lockdown period respectively. The concurrent presence of fatigue, cough, and diarrhea in children, in absence of sore throat/earache and abnormal skin signs, represents the maximum risk level of having a suspected/confirmed case of COVID-19 at home.

**Conclusions:** The percentage of children presenting COVID-19 like symptoms at home has been remarkable also during the total lockdown period. The present study identified a pattern of symptoms which could help, in a cost-effective perspective, PCPs in daily clinical practice to define priorities in addressing children to the proper diagnostic procedure.

## Introduction

Much information has been learned about the infection of SARS-CoV-2 and coronavirus disease (COVID-19), and much more has still to be learned, especially in children [1]. Indeed COVID-19 is rare in the pediatric population, both in China and in Europe [2-4]. In Italy cumulative incidence COVID-19 is as low as 0.42/1000 in the first decade of life and peaks to 24.6/1000 in people aged 90 years and over [5]. Moreover, COVID-19 in pediatric age is usually milder and with better prognosis than in adults and older people, likely due to both exposure and host factors [6,7]. This was confirmed worldwide and also in Italy [8]. Fatality rate is 0.2% in the first decade of life and becomes > 30% in people aged 80 years and over [5].

Several mechanisms have been proposed to explain the favorable pattern of COVID-19 in children. SARS-CoV-2 is able to penetrate cells thanks to the binding to angiotensin-converting enzyme 2 (ACE2), and then to downregulate it. ACE2 cleaves angiotensin 2 (with a pro-inflammation and pro-fibrosis role) to angiotensin (1-7) form (which is active in anti-inflammation pathway) [9]. In children, ACE2 binding domain may be immature and dysfunctional, thus preventing optimal infectious process, but at the same time ACE2 levels are higher and may provide higher concentration of angiotensin (1-7) heptapeptide [10-12]. Furthermore, children show stronger innate immunity secondary to live-vaccines and high incidence of respiratory virus infections, whereas immune-senescence is found in elderly [10]. Finally, children may have had fewer risk contacts during the spread of the SARS-CoV-2 infection than adults and elderly people because of early closing of schools [13].

Infection in children may be asymptomatic or may occur with a pool of typical symptoms all over the world: acute upper respiratory tract symptoms including fever, fatigue, myalgia, cough, sore throat, pharyngeal erythema, runny nose, earache, and sneezing have been the first associated with mild SARS-COV-2 infection in pediatric age. Later digestive symptoms, such as nausea, vomiting, abdominal pain, and diarrhea were observed in COVID-19 positive children. Cutaneous manifestations can be additionally found in confirmed cases. Moderate-to-severe infections are characterized by pneumonia, tachypnea, or tachycardia [14-18]. In our study, these typical COVID-19 symptoms in absence of a firm diagnosis were defined as “COVID-19 like symptoms”.

Since 1978 the Italian National Health Service (NHS) has provided Pediatric Primary Care (PPC) to children through the use of Primary Care Pediatricians (PCPs), (Law 833/1978 and subsequent ones concerning the National Health Service) [19]. The organization of PPC in Italy is unique in Europe as even the first line management of children is handed over to the PCPs till the end of the developmental age. The task of Italian PCPs is to answer the psychosocial and developmental needs of children, assisting families in promoting their children’s health and well-being [20]. Of note, the pediatric healthcare global cost for the National Health fund is 0.2–0.3% of overall budget [21].

During this outbreak it was quite tricky when a family with a child with acute illness (fever, respiratory or gastrointestinal symptoms, and cutaneous signs) reach out PCP for a diagnosis: according to precautionary principles, European recommendation is to visit this child with all the necessary personal protective equipment (PPE) [22]. It must be underlined that PCPs in Italy were not allowed to directly perform the nasopharyngeal swab, and request for it to be performed by responsible healthcare personnel could be possible only if a confirmed case is found in the child family, or no clinical diagnosis was feasible.

Primary aim of our study was to find out whether outpatient children have developed COVID-19 like symptoms during Italian lockdown and social distancing, and to explore the impact of COVID-19 like disorders on PPC system. Secondary aim was to provide PCPs useful insights to be used in clinical practice during the difficult clinical differential diagnosis process in the COVID-19 era.

## Materials and methods

### Study population

From the 4^th^ of March to the 23^rd^ of May 2020, during the Italian lockdown period, an online survey with anonymous questionnaires was submitted to 50 PCPs, assisting 47,500 children aged 0-16 years, from 7 different Italian regions. COVID-19 like symptoms were identified in Literature as the most common in COVID-19 positive children [14,15,17,23,24], and defined as the presence of one of the following: fever, fatigue, rhinitis, sore throat or earache, cough, nausea or vomiting, diarrhea, or abnormal skin sign. PCPs completed the questionnaires for each child with COVID-19 like symptoms addressing their surgeries. During the first part of the outbreak a long latency was present between naso-pharyngeal swab request and results, thus swab results, if performed, were not collected in the survey. Baseline clinical data were collected in the questionnaires. The presence of confirmed or suspected cases in children’s families was recorded according to the WHO Global Surveillance for human infection with COVID-19 definitions [25].

Italian regions were classified as low, medium or high-risk areas based on the 26^th^ of May COVID-19 update, issued by the Italian National Institute of Health [26]. In detail, Calabria and Sardinia were low risk areas, Lazio medium risk area, Emilia-Romagna, Liguria, Lombardy and Piedmont high-risk areas. The observation period was divided in: total lockdown, from the 4^th^ of March till the 3^rd^ of May, and partial lockdown, from the 4^th^ till the 23^rd^ of May.

### Statistical analyses

Significance of the association between area of residence, gender, age class, symptoms and presence of suspected/confirmed cases in the family was evaluated by Fisher’s exact test. Multivariable analysis was accomplished by a logistic regression model, where suspected/confirmed cases in the family (0=absent, 1=present) was the response variable, and area of residence (0=low/medium risk, 1= high risk), gender, age (<1, 1-5, 6-10, 11-16 years) and symptoms were the potential determinants. Results were reported as Odds Ratios (ORs), and p values were computed by the Wald test. Goodness-of-fit was evaluated by the Hosmer-Lemeshow test. Coefficients derived from logistic regression were used to estimate the risk of suspected/confirmed cases in specific subgroups. Prevalence of symptoms in our study population was compared with the one reported by Parri et al. considering a cohort of 100 Italian children assessed in 17 Pediatric Hospitals and positive to SARS-CoV-2 [27].

## Results

2,300 children, representing the 4.8% (95% CI 4.7-5.0%) of our whole pediatric population, visited PCPs for COVID-19 like symptoms. 1,464 children were visited from the 4^th^ of March till the 3^rd^ of May, and 836 from the 4^th^ till the 23^rd^ of May, so that the average number of visits per working day was 35.7 during the total lockdown and 55.7 during the partial lockdown.

Males (n=1,242, 54.0%) were more prevalent than females (n=1,058, 46%), and median age was 4 years (range 0-16 years). Age distribution significantly differed between the Italian hospital series recently published [27] and the PPC survey (present study), as the former mostly comprised children in the first year of life or adolescents aged 11-18 years, while the latter mostly recruited children aged 1-5 years (53.6%) or 6-10 years (26.1%) (Table 1). Comorbidities affected 27% of hospitalized children, while they were rare in the PPC series. On the contrary, fever, fatigue and skin abnormalities were more frequently reported in children visited in PPC than in the hospital (Table 1).

**Table 1.** Symptoms of COVID-19 + children (prevalence) in a hospital series compared with symptoms of children attending PPC in Italy (modified from [27]).

|  | **Hospital series [27]**<br>**(n=100)** | **Pediatric Primary Care**<br>**survey (n=2300)** | **p value\*** |
| --- | --- | --- | --- |
| Males (%) | 57 (57%) | 1242 (54.0%) | 0.609 |
| Age class (years) |  |  | **<0.001** |
| <1 | 40 (**40%**) | 153 (6.7%) |  |
| 1-5 | 15 (15%) | 1233 (**53.6%**) |  |
| 6-10 | 21 (21%) | 601 (**26.1%**) |  |
| >10** | 24 (**24%**) | 313 (13.6%) |  |
| Comorbidities | 27 (27%) | 55 (2.4%) | **< 0.001** |
| Fever | 54 (54) | 1479 (**64.3**) | **0.043** |
| Cough | 44 (44) | 871 (37.9) | 0.247 |
| Rhinorrhea | 22 (22) | 571 (24.8) | 0.556 |
| Shortness of breath | 11 (11) | --- |  |
| Drowsiness | 11 (11) | --- |  |
| Nausea or vomiting | 10 (10) | 294 (12.8) | 0.538 |
| Diarrhea | 9 (9) | 380 (16.5) | 0.051 |
| Fatigue | 9 (9) | 472 (**20.5**) | **0.003** |
| Abnormal skin signs | 3 (3) | 283 (**11.8**) | **0.004** |

\*p values were computed by Fisher's exact test.
\*\*10-18 years in the hospital series, 10-16 years in the PPC survey.

As expected, the proportion of children living with suspected and confirmed cases was significantly greater in high than medium/low risk areas and during total than partial lockdown (p<0.001). The risk of suspected/confirmed cases in the family was affected by neither gender nor the presence of comorbidities, and increased with increasing age (Table 2).

**Table 2.** COVID-19 infection in the children’s family as a function of gender, age, residence, time of the survey.

|  | N | No infection<br>(n=1892) | Suspected<br>(n=225) | Confirmed<br>(n=183) | P value* |
| --- | --- | --- | --- | --- | --- |
| Residence |  |  |  |  | **<0.001** |
| Low risk area | 927 | 868 (**93.6**) | 53 (5.7) | 6 (0.6) |  |
| Medium risk area | 153 | 145 (**94.8**) | 3 (2.0) | 5 (3.3) |  |
| High risk area | 1220 | 879 (72.0) | 169 (**13.9**) | 172 (**14.1**) |  |
| Time of the survey |  |  |  |  | **<0.001** |
| Total lockdown | 1464 | 1141 (77.9) | 181 (**12.4**) | 142 (**9.7**) |  |
| Partial lockdown | 836 | 751 (**89.8**) | 44 (5.3) | 41 (4.9) |  |
| Gender |  |  |  |  | 0.113 |
| Males | 1242 | 1011 (81.4) | 136 (11.0) | 95 (7.6) |  |
| Females | 1058 | 881 (83.3) | 89 (8.4) | 88 (8.3) |  |
| Age class (years) |  |  |  |  | **<0.001** |
| <1 | 153 | 139 (**90.8**) | 8 (5.2) | 6 (3.9) |  |
| 1-5 | 1233 | 1062 (**86.1**) | 100 (8.1) | 71 (5.8) |  |
| 6-10 | 601 | 475 (79.0) | 65 (**10.8**) | 61 (**10.1**) |  |
| 11-16 | 313 | 216 (69.0) | 52 (**16.6**) | 45 (**14.4**) |  |
| Comorbidities |  |  |  |  | 0.493 |
| No | 2245 | 1843 (97.4) | 222 (98.7) | 180 (98.4) |  |
| Yes | 55 | 49 (2.6) | 3 (1.3) | 3 (1.6) |  |

Significance differences are highlighted in bold.

Children living in a family with confirmed COVID-19 cases had a higher prevalence of diarrhea and fatigue, and a lower prevalence of sore throat/earache and abnormal skin signs than children with no family cases. Accordingly, also the prevalence of nausea and vomiting tended to be the highest among children with confirmed cases, but the difference was not significant. Fever and cough were more common among children living in a family with suspected COVID-19 cases. A possible explanation could be that fever and cough are more common in the early phase of COVID-19 infection, while diarrhea and fatigue prevail later on (Table 3).

**Table 3.** Children with symptoms (percent) as a function of COVID-19 infection in the family.

|  | No infection<br>(n=1892) | Suspected<br>(n=225) | COVID-19<br>positive<br>(n=183) | P value* |
| --- | --- | --- | --- | --- |
| Fever | 1,217 (**64.3**) | 158 (**70.2**) | 104 (56.8) | **0.020** |
| Fatigue | 327 (17.3) | 76 (**33.8**) | 69 (**37.7**) | **<0.001** |
| Rhinitis | 471 (24.9) | 57 (25.3) | 43 (23.5) | 0.908 |
| Sore throat / earache | 694 (**36.7**) | 56 (24.9) | 38 (20.8) | **<0.001** |
| Cough | 667 (35.3) | 128 (**56.9**) | 76 (41.5) | **<0.001** |
| Nausea or vomiting | 236 (12.5) | 27 (12.0) | 31 (16.9) | 0.215 |
| Diarrhea | 286 (15.1) | 46 (**20.4**) | 48 (**26.2**) | **<0.001** |
| Abnormal skin signs | 251 (**13.3**) | 18 (8.0) | 14 (7.7) | **0.009** |

\*P value was computed by the Fisher's exact test.

In multivariable analysis, the risk to have suspected/confirmed cases at home was five-fold higher in high than low/medium risk areas, increased with increasing age, and was not affected by gender. Among symptoms, fatigue, cough and diarrhea were suggestive of family suspected/confirmed cases, while sore throat/earache and abnormal skin symptoms were associated with no cases at home (Table 4).

**Table 4.** Risk factors for the presence of suspected/confirmed case(s) in the family.

|  | OR (95% CI) | P value* |
| --- | --- | --- |
| Residence (high vs low risk area) | **5.32 (4.00-7.08)** | **<0.001** |
| Gender (Females vs Males) | 0.98 (0.77-1.24) | 0.852 |
| Age (years) |  |  |
| <1 | 1 (reference) |  |
| 1-5 | **1.92 (1.06-3.49)** | **0.033** |
| 6-10 | **2.77 (1.50-5.14)** | **0.001** |
| 11-16 | **3.82 (2.02-7.21)** | **<0.001** |
| Symptoms (yes vs no) |  |  |
| Fever | 0.97 (0.75-1.26) | 0.835 |
| Fatigue | **2.10 (1.60-2.74)** | **<0.001** |
| Rhinitis | 0.96 (0.72-1.29) | 0.799 |
| Sore throat / earache | **0.50 (0.38-0.67)** | **<0.001** |
| Cough | **1.97 (1.51-2.56)** | **<0.001** |
| Nausea or vomiting | 0.83 (0.57-1.20) | 0.312 |
| Diarrhea | **1.68 (1.21-2.33)** | **0.002** |
| Abnormal skin signs | **0.56 (0.37-0.87)** | **0.009** |

\*P values was computed by the Wald test
Odds Ratios (OR) and the corresponding 95% Confidence Interval (95% CI) was compute by a logistic regression model.

The risk of living with a suspected/confirmed case was quantified for high risk areas (Figure 1) and low/medium risk areas (Figure 2) through the coefficients estimated by the logistic regression model. For instance, baseline risk of having a suspected/confirmed case in the family was about 30% in the children aged 6-10 years from high risk areas, it increased to about 40% in children with fatigue, or cough or diarrhea, to about 50% in the presence of 2 of these symptoms, and peaked to 62.6% when the 3 symptoms were simultaneously present. On the contrary, this probability decreased to about 20% in children with sore throat/earache or skin symptoms, and further to 14.3% in the presence of both symptoms. The corresponding figures were much lower for children aged 6-10 years living in low/medium risk areas. The baseline risk of having suspected/confirmed cases in the family was 8.1%, it increased to 11-13% in children with fatigue or cough or diarrhoea, to 16-18% in the presence of 2 of these symptoms, and peaked to 24.6% when the 3 symptoms were simultaneously present. The risk became very low (5%) in children with sore throat/earache or abnormal skin symptoms, and even lower (3%) in the presence of both symptoms.

**Fig 1.**
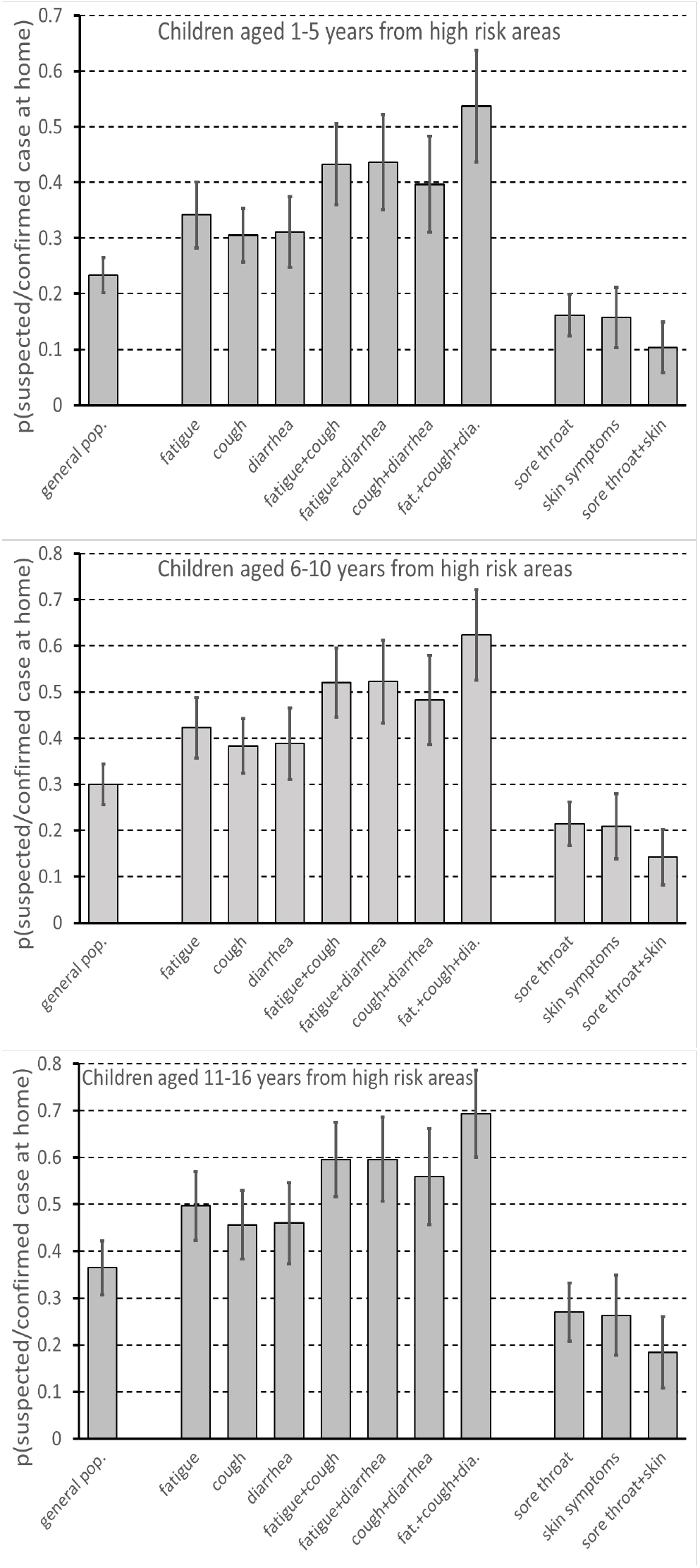
Probability of having a suspected/confirmed case in the family as a function of symptoms, which were significant predictors in the multivariable logistic model. Estimates refer to children aged 1-5 years (upper panel), 6-10 years (middle panel), 11-16 years (lower panel) living in HIGH risk areas. Columns are probabilities, bars are 95% confidence intervals. Sore throat = sore throat/earache.

**Fig 2.**
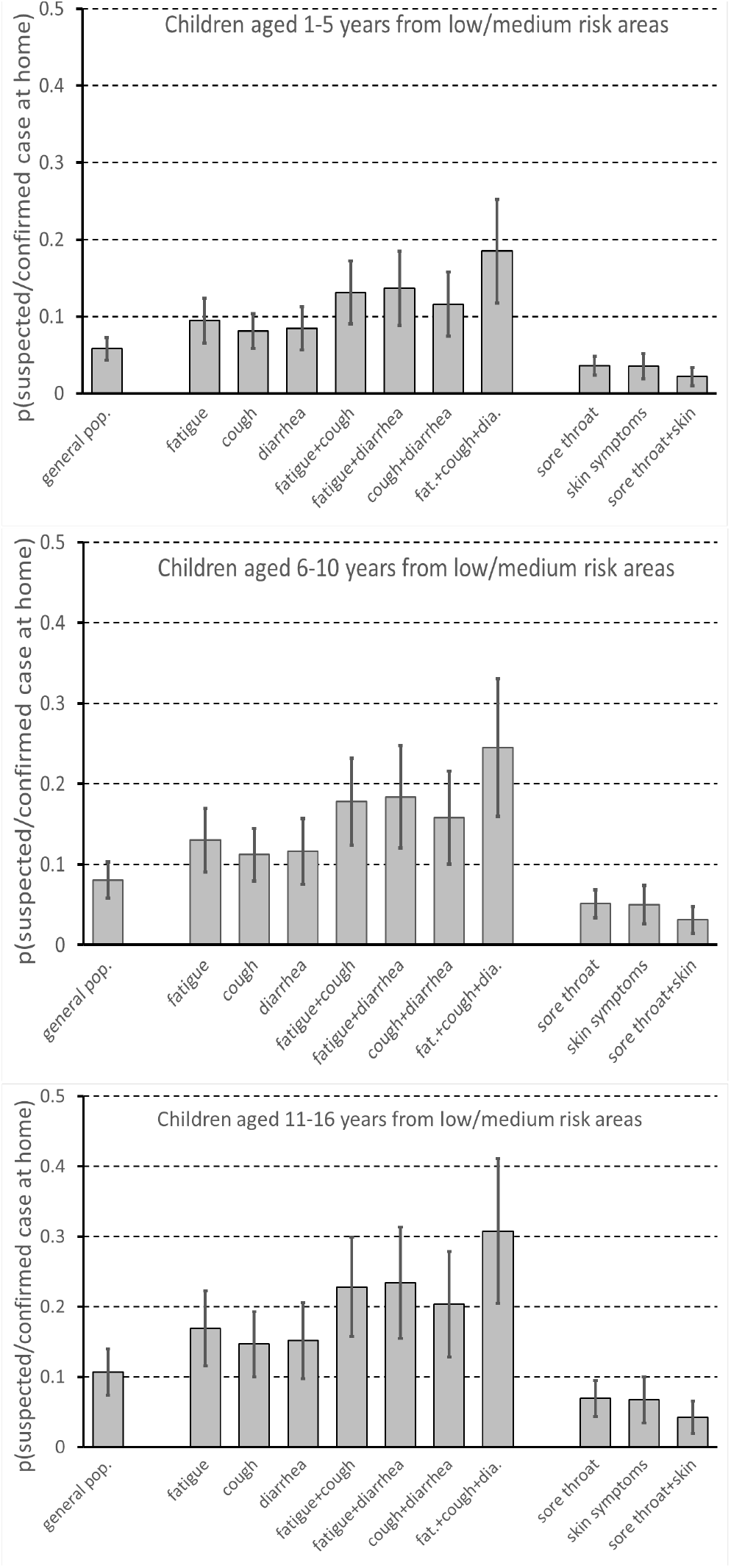
Probability of having a suspected/confirmed case in the family as a function of symptoms, which were significant predictors in the multivariable logistic model. Estimates refer to children aged 1-5 years (upper panel), 6-10 years (middle panel), 11-16 years (lower panel) living in LOW/MEDIUM risk areas. Columns are probabilities, bars are 95% confidence intervals. Sore throat = sore throat/earache.

## Discussion

This is the first survey held in Italy with the PCPs, addressing the pattern of pediatric surgeries access during the complete lockdown and for 20 days thereafter. Nowadays the actual number of COVID-19 cases in Italy is still unknown, and probably underestimated, especially in the pediatric population [5]. Of note the percentage of children presenting COVID-19 like symptoms at home has been remarkable during the lockdown period (4.8%). This was unexpected, as children are less likely to get sick when the schools are closed. The proportion of children with suspected/confirmed cases in the family more than halved from the total to the partial lockdown period, reflecting the decrease in COVID19 incidence in Italy [5]. However, access to pediatric surgeries increased from the total to the partial lockdown period from 35.7 to 55.7 visits/working day, and this probably reflects increased family mobility and persistent parents’ anxiety. The risk of having suspected/confirmed cases at home was 5-fold larger in children living in high than low-medium risk areas, and increased with increasing age, in agreement with Italian epidemiological data. Indeed, the cumulative incidence of COVID-19 by the 23^rd^ of June 2020 was 0.42/1000 in children in the first decade of life and increased to 0.67/1000 in the second decade. In the whole population cumulative incidence ranged from 0.60/1000 in Calabria to 9.25/1000 in Lombardy [5].

Interestingly, the prevalence of major COVID-like symptoms in Italy is intriguingly similar between the present study on children addressing pediatric surgeries, who did not perform any swab, and the study by Parri et al. [27] on children admitted to the hospital with positive swab. This similarity suggests that prevalence of COVID-19 could be actually underestimated in Italian outpatient children. Anyway, PCPs were not enabled to do the differential diagnosis between COVID-19 and other disorders as they lack the necessary diagnostic tools.

The main practical finding of our study is that the concurrent presence of fatigue, cough, and diarrhea in children, in absence of sore throat/earache and abnormal skin signs, represents the maximum risk level of having a suspected/confirmed case of COVID-19 at home, which was assumed as a proxy of COVID-19 in the present study. A dose-response association with symptoms having a suspected/confirmed case at home emerged, as the risk increased from children without fatigue, cough and diarrhea to children with only 1 symptom and further to children with 2 or 3 symptoms. On the other hand, a child who presents both sore throat/earache and cutaneous manifestations and no fatigue, cough, or diarrhea, was less likely to have a suspected/confirmed case at home, and hence to be infected with SARS-CoV-2.

Given the importance to early identify COVID-19 positive outpatient children, our risk score could be a useful instrument in PCPs clinical practice to define priorities in addressing children to the proper diagnostic procedure, even if they have not affected familiars. As suggested by the Federal Office of Public Health (UFSP) of the Swiss Confederation [28], not all the outpatient children with mild COVID-19 like symptoms need to be tested for COVID-19, and the final decision is up to PCPs and parents.

To our knowledge, this is the first survey held in Italy with the PCPs, addressing the pattern of pediatric surgeries access during the complete lockdown and for 20 days thereafter. Moreover, the survey considered 2,300 children from 7 Italian regions. The main limitation of this study was the lack of nasopharyngeal swab as a diagnostic tool for SARS-CoV-2 infection. Of note, this limitation was not peculiar of the present study but affected the entire primary care setting in Italy, for both children and adults. To circumvent this limitation the presence of suspected/confirmed case of COVID-19 at home was assumed as a proxy of COVID-19.

## Conclusions

The percentage of children presenting COVID-19 like symptoms at home has been remarkable also during the total lockdown period. COVID-19 pandemic is not over and could outbreak again in autumn. Health care resources are limited, so it is not realistic to perform a nasopharyngeal swab in all symptomatic children. The present study identified a pattern of symptoms which could help, in a cost-effective perspective, PCPs in daily clinical practice, to define priorities in addressing children to the proper diagnostic procedure.

## Data Availability

Data supporting the findings of this study are available from the corresponding author GM on request.

## Acknowledgments

We warmly thank all the membership of the Research Collaborative Group of Primary Care Pediatricians: Giulia Franchi, Marisa Gramegna, Giorgio Lepre, Cosimo Claudio Muià, Mirella Nigro, Luisella Prete, Karim Primon, Emanuela Pilade, Maria Adele Zoia, Claudia Piasenti, Daniela Piana, Fabrizio Fraioli, Mario Moi, Cristina Perrera, Simonetta Serra, Tiziana Casti, Paolo Rosas, Miriam Concas, Pietro Maria Di Giovanni, Pierangela Mussinu, Giuseppe Cera, Antonio Pala, Bruna Moi, Sabrina Pilia, Licia Mocci, Cosimo Lanza, Marcello Semprini, Antonella Lavagetto, Anna Timitilli, Carola Malfatti, Roberto Bezzi, Livio Fattorini, Franca Passeri, Donato Orrù, Cristian Noto, Paola Pescosolido, Patrizio Veronelli, Nadia Chiapello, Maria Grazia Giuliani, Norberto Porta, Giuseppe Squazzini, Lorenzo Parodi, Fulvia Gatto, Giovanna Levato, and Giulietta Manca.

## Notes

### Competing Interest Statement

The authors have declared no competing interest.

### Funding Statement

The authors received no financial support for the research, authorship, and/or publication of this article.

### Author Declarations

Comitato Etico regione Liguria - Ospedale San Martino di Genova. (protocol number 10759 - study number 375/2020)

